# Understanding the Impacts and Perceptions of Alcohol Use in Northern Tanzania: A Mixed-Methods Analysis

**DOI:** 10.1101/2023.09.11.23295395

**Authors:** Alena Pauley, Madeline Metcalf, Mia Buono, Kirstin West, Sharla Rent, William Nkenguye, Yvonne Sawe, Mariana Mikindo, Joseph Kilasara, Bariki Mchome, Blandina T. Mmbaga, João Ricardo Nickenig Vissoci, Catherine A. Staton

## Abstract

**Background:** Worldwide, alcohol is a leading risk factor for death and disability. Tanzania has particularly high rates of consumption and few resources dedicated to minimizing alcohol-related harm. Ongoing policy efforts are hampered by dynamic sociocultural, economic, and regulatory factors contributing to alcohol consumption. Through the voices of Kilimanjaro Christian Medical Center (KCMC) patients and a gender-focused lens, the goal of this project was to investigate community perceptions surrounding alcohol and the impact of its use in this region.

**Methods:** This was a mixed-methods study conducted at KCMC between October 2021 and May 2022. 678 adult (≥18 years old) Kiswahili-speaking patients who presented to KCMC’s Emergency Department (ED) or Reproductive Health Clinic (RHC) were enrolled through systematic random sampling to participate in quantitative surveys. Nineteen participants were selected for in-depth interviews (IDIs) through purposeful sampling. The impact and perceptions of alcohol use were measured through Drinkers’ Inventory of Consequences (DrInC) scores analyzed in R Studio through descriptive proportions, and IDI responses explored through a grounded theory approach utilizing both inductive and deductive coding methodologies.

**Results:** ED men were found to have the highest average [SD] DrInC scores (16.4 [19.6]), followed by ED women (9.11 [13.1]), and RHC women (5.47 [9.33]), with higher scores indicating greater perceived consequences. Participants noted alcohol has both perceived advantages and clear harms within their community. Increased conflict, long-term health outcomes, financial instability, stigma, and sexual assault were seen as negative consequences. Benefits were primarily identified for men and included upholding cultural practices, economic growth, and social unity. Physical and financial harm from alcohol impacted both genders, however, alcohol-related stigma and sexual assault were found to disproportionately affect women.

**Conclusion:** Our findings suggest that perceptions around drinking and alcohol’s social and physical consequences differ significantly by gender. To effectively minimize local alcohol-related harm, future alcohol-focused research and policy efforts should consider the distinct impacts alcohol has between genders.

## Introduction

Alcohol consumption has been implicated as a leading cause of harm, disability, and death worldwide (1). The harmful use of alcohol is a growing and avoidable cause of premature disability and death, each year causing 5.1% of all disability-adjusted life years (DALYs) and 3 million deaths globally (1). Alcohol use has been linked to over 200 medical conditions, including digestive diseases, infectious diseases, cardiovascular disease, certain cancers, and injuries(2–4), and has also been associated with poor employment outcomes (5), individual and societal economic harm (4,6–8), social stigma (9,10), and sexual violence (11–13).

Although a global problem, alcohol use is increasing especially rapidly in low and middle-income countries (14–16). For example, alcohol use has become the leading risk factor for disease burden in many sub-Saharan African countries (17,18). As low-income populations typically have fewer resources, financial savings, and poorer access to health care services, alcohol-related harm in these contexts is more devastating both physically and financially (1,19–21). While the harms of alcohol intake are significant, its use remains endemic and has been ingrained in cultural and social practices in different populations across the world (22). This holds true in Tanzania, where the cultural and social practices of alcohol use, such as normalization of use from a young age and use as a social currency, support larger quantities of consumption (23).

Adding greater complexity to alcohol’s impact on individuals and communities, alcohol consumption has been shown to differ in populations according to cultural norms, regulation, age, socio-economic status, etc (24). To address the factors influencing alcohol consumption, the World Health Organization (WHO) created the Conceptual Model Of Alcohol Consumption and Health Outcomes. The model aims to decrease the morbidity and mortality that follow harmful alcohol use patterns and demonstrates that understanding an individual country’s sociocultural context is imperative to alter alcohol use behaviors at an individual and country level (25).

Moshi, located near Mount Kilimanjaro National Park in Tanzania, has seen exceptionally high rates of alcohol use, 2.5 times higher than in nearby regions (26,27). Previous studies have shown that 60% of Moshi residents frequently consume alcohol, and 23% suffer from alcohol use disorder (28). In this location, alcohol consumption has also been linked here to greater stigma (29), unsafe sexual behaviors (30,31), and an increased prevalence of injuries (32). While alcohol-related harms have been identified in Moshi, current literature specifying how community members view the harms and benefits of alcohol holistically is sparse.

Previous work from our team has examined stigma (29,33), perceptions of alcohol use (23), the prevalence of alcohol-related injuries upon Emergency Department (ED) arrival (32), and developing and implementing an intervention to reduce alcohol misuse for the ED injury patient population (34,35). Building off of this research, a greater understanding of the specific societal perceptions of alcohol will allow for a clearer picture of its role within the Moshi community. This, in turn, can help inform and shape future alcohol-reduction interventions and research initiatives to be more effective and successful in decreasing alcohol-related harm in this setting (36). This process of development can also serve as a model for population-specific harm reduction interventions in other settings. Thus, through the voices of patients at the Kilimanjaro Christian Medical Center (KCMC), this study aims to fill this gap by investigating the negative and positive ways that alcohol use impacts individuals and the broader Moshi community along with perceptions of alcohol use. Factors underlying differential expectations of use will also be explored.

## Methods

### Study Design

This is a secondary analysis of a mixed-methods study on gender differences in alcohol use among KCMC’s Emergency Department (ED) and Reproductive Health Center (RHC) patients in Moshi, Tanzania. The ED was chosen as one of the study sites to better understand risky alcohol use behaviors given the correlation between excessive alcohol use and injuries that prompt ED visits (32,37,38). The RHC, on the other hand, was selected because of its primarily female patient population, which facilitated a richer, gynocentric perspective on alcohol use.

Data for this study were collected between October 2021 and May 2022 and consisted of quantitative surveys and qualitative in-depth interviews (IDIs). During this time, 678 patients were enrolled, but only 655 participants fully completed survey questionnaires. These participants were enrolled through a systematic random sampling strategy to ensure a balanced representation of each patient population. Of the pool of individuals already participating in this study, 19 were chosen via purposive sampling for IDIs. A full description of the methods for this project is noted elsewhere (39).

This current analysis seeks to understand 1) the impacts alcohol use has on patients themselves and 2) how they perceive alcohol to affect their community. In answering these questions, a quantitative tool that assesses an individual’s alcohol-related consequences, the Drinkers Inventory of Consequences (DrInC), was used in addition to a qualitative exploration of the perceived harms and benefits of alcohol use.

### Study Sample

All participants were recruited from KCMC’s RHC or ED. Those enrolled met the following inclusion criteria: 1) fluency in Kiswahili, 2) ability to provide informed consent, 3) aged 18 or older, and 4) not a prisoner.

### Quantitative Data

#### Data Collection

KCMC ED and RHC patients were approached for participation in this study by a same-gender, Kiswahili-speaking, Tanzanian research assistant trained in Good Clinical Practices and study procedures. A systematic random sampling strategy was used to guide enrollment, where every third patient was approached based on ED and RHC intake registries, with the exception of ED females. Given that KCMC’s ED patient population is heavily male, to maintain enrollment goals, every female who presented to the ED was approached.

After the research assistants ensured patients were medically stable, they approached patients in a quiet, private location and provided an introduction to the study. All patients had the opportunity to decline participation, but if willing to proceed, an in-depth discussion of the risks and benefits of study participation was provided. If agreeing, written consent was obtained, and survey questions were delivered. All questions were asked orally to encourage participation from all literacy levels.

#### Instruments

The DrInC was used to quantitatively assess the impact alcohol use had on an individual. DrInC, which had been cross-culturally adapted and clinically tested previously at KCMC (40), is a 50-item survey tool consisting of yes or no questions designed to assess alcohol-related consequences across five different domains (41,42). These include physical, social responsibility, interpersonal, intrapersonal, and impulse control (Table 1). DrInC scores range from 0 to 50, with higher scores overall indicating more affirmative responses to various alcohol-related items as shown in Table 1 below. Of note, while DrInC helps to quantitatively illustrate the range of alcohol-related consequences one has experienced, it does not measure the severity or frequency of these consequences. Further, no particular clinically relevant cut-off score for this tool exists.

**Table 1:**
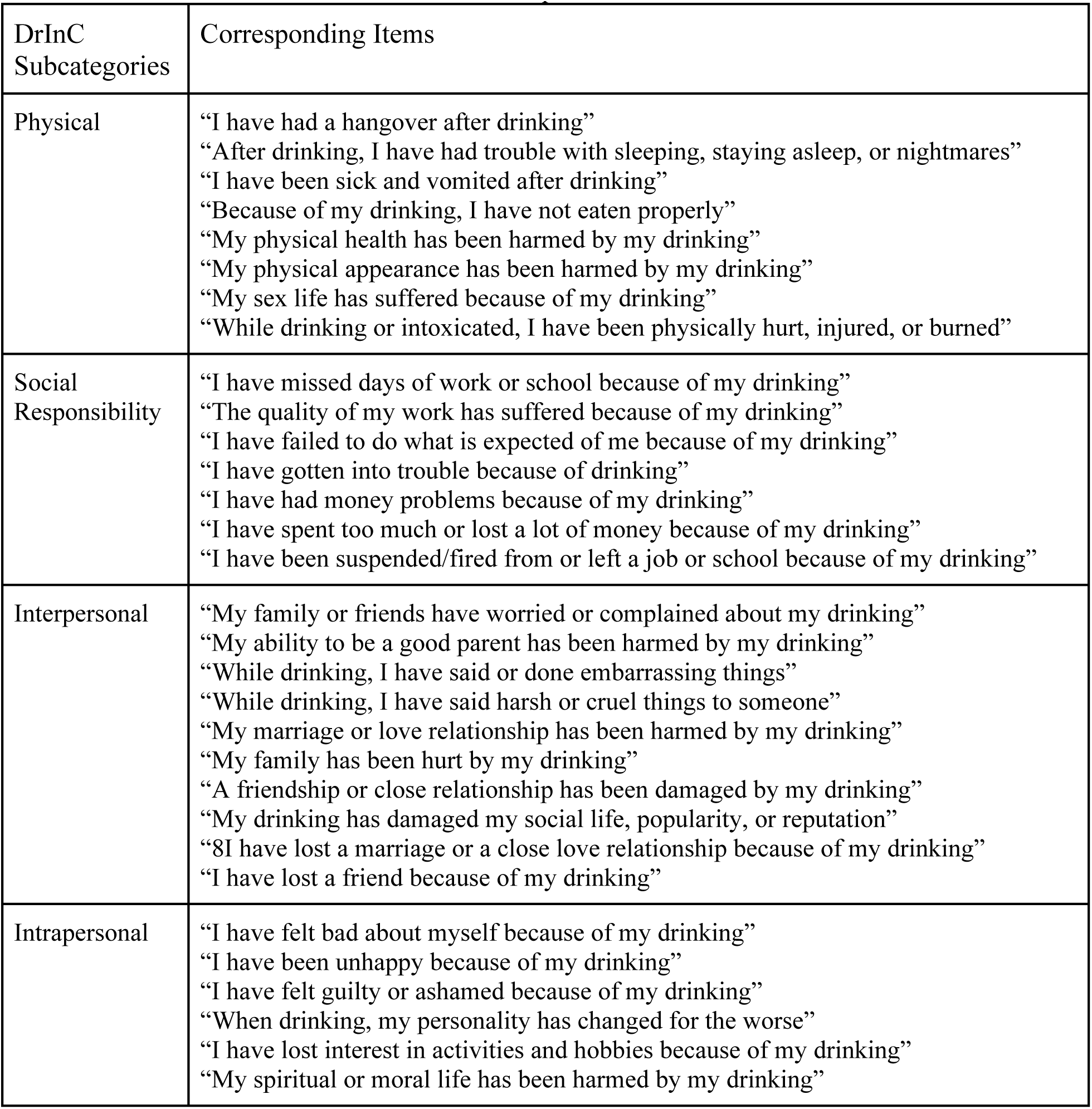
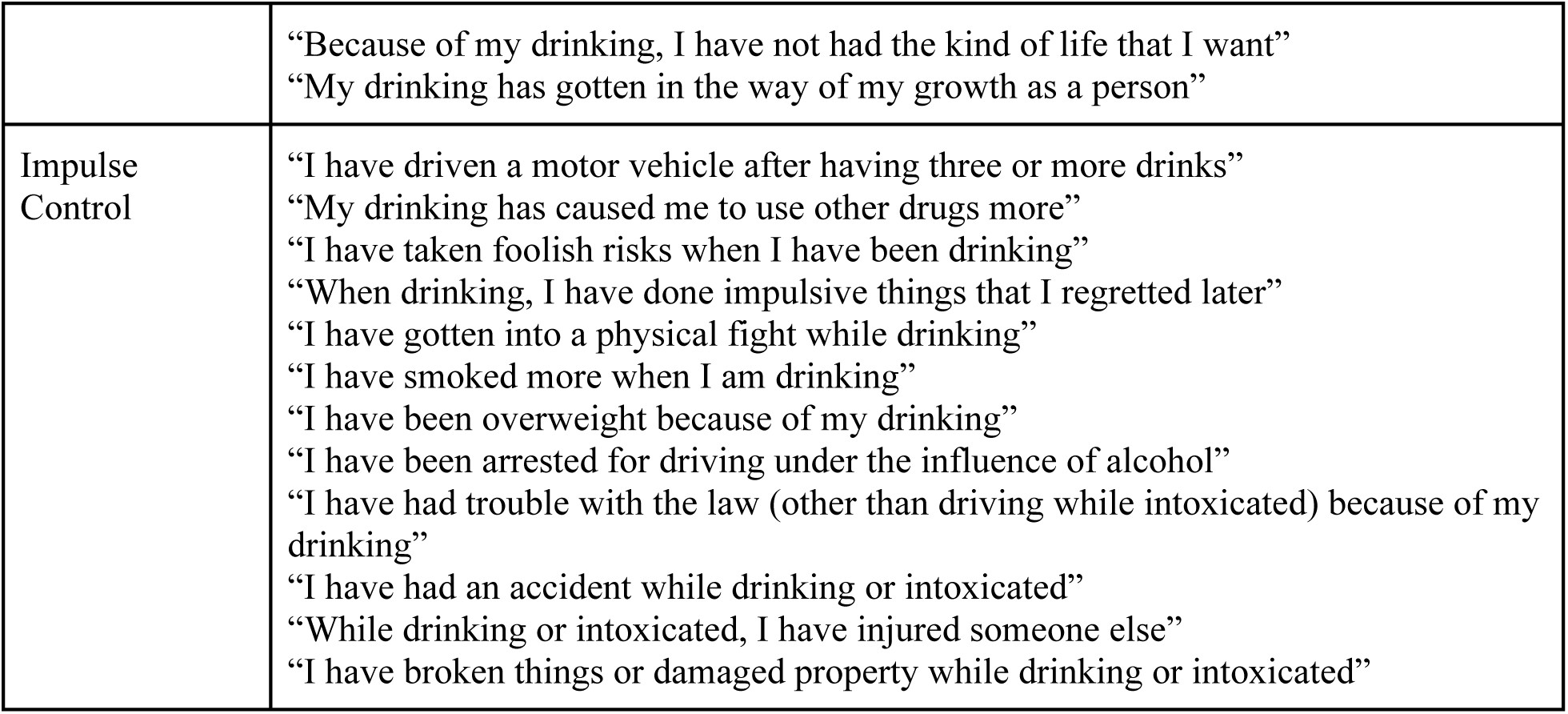
Consequence Domains and Corresponding Items of the Drinkers’ Inventory of Consequences.

#### Analysis

Overall and category specific DrInC scores for the three patient populations were assessed through descriptive frequencies and proportions. DrInC scores were also analyzed in relation to patients’ alcohol intake to identify if an association between increased alcohol consumption and more consequences existed. Alcohol consumption was measured by the Alcohol Use Disorder Identification Test (AUDIT), a clinical tool used to identify patients with unhealthy drinking behaviors that was administered at the same time as the DrInC survey. AUDIT scores range from 0 to 40, with higher scores representing more hazardous alcohol consumption and scores of 8 or greater clinically indicative of risk for adverse alcohol-related health effects both globally and in the study setting (43–46). The relationship between patient DrInC and AUDIT scores was measured through a linear regression analysis, the independent variable being DrInC scores and the dependent variable being AUDIT scores. Incomplete surveys were still included in analyses. All quantitative data were analyzed in R Studio.

### Qualitative Data

#### Data Collection

Nineteen out of the 678 patients enrolled were selected to participate in IDIs. Participants were chosen through purposive sampling to be balanced by gender, encompass diverse demographic backgrounds in terms of age, religion, income, profession, and marital status, and hold varying degrees of personal alcohol consumption. Additionally, the research team strove to have multiple unique experiences with alcohol use represented in the final data set, from patients who only reported neutral or positive past experiences with alcohol, used to drink but are now abstinent, have a close friend or relative that drinks heavily, or suffered significant negative consequences from alcohol. Interviewee characteristics were re-reviewed after two subsequent IDIs, and any needed changes were implemented when selecting the next IDI candidate.

These patients were approached for IDI participation at the conclusion of their initial survey or in a later phone call. If the patient expressed interest in participating, the same-gendered research assistant who had originally enrolled them set up a time to meet and conducted the interview with them in a private room at KCMC. All interviews were conducted in Kiswahili using a semi-structured interview guide and were recorded on a handheld audio recorder. The research assistant conducting the interview asked probing follow-up questions when information was unclear, conflicted with previous statements, or when a notable story warranted further explanation. IDI participants were given a small stipend to reimburse their travels and were offered a snack and drink midway through.

#### Analysis

An applied thematic, grounded theory approach was used to inductively and deductively analyze all qualitative data. Iterative development of the initial codebook as led by Tanzanian and US team members, facilitated the creation of content memos per each emerging theme and code. These content memos then served as the foundation for the final analysis after discussion and feedback with the full research team.

#### Ethics Statement

Ethical approval for this study was obtained from the Duke University Institutional Review Board, the Kilimanjaro Christian Medical University College Ethical Review Board, and the Tanzanian National Institute of Medical Research prior to any data collection. Personal health information was employed in screening and enrollment procedures, but was de-identified when collected, stored, and analyzed and shared only via a data share agreement.

## Results

### Quantitative

Six-hundred and seventy-eight patients started but only 655 patients completed our survey questionnaire, where ED men were found to spend the most money on alcohol per week, drink the most frequently and in the largest quantities, and have the highest average AUDIT scores as compared to women ED and RHC patients (39). In comparing alcohol consumption with alcohol-related consequences, AUDIT and DrInC scores were found to be linearly associated across all three patient populations (Figure 1). This association was strongest for women ED patients (R² = 0.76; p<0.001) and weakest for men ED patients (R² = 0.65; p<0.001) and women RHC patients (R² = 0.53; p<0.001).

**Figure 1:**
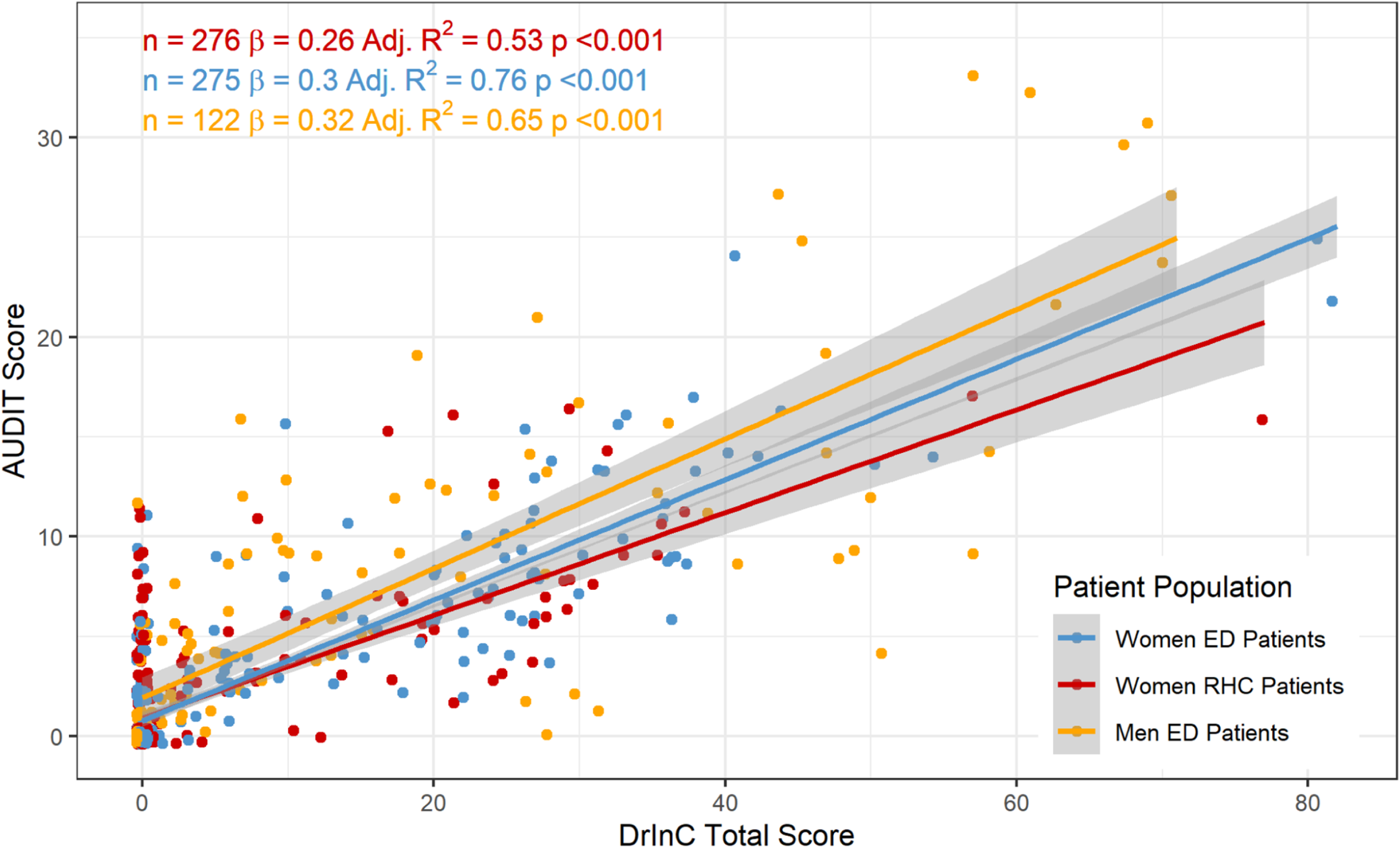
Association of AUDIT and DrInC Scores across Patient Populations

For alcohol-related consequences, ED men were found to have the highest average [SD] DrInC scores (16.4 [19.6]), followed by ED women (9.11 [13.1]), and RHC women (5.47 [9.33]) (Table 2). This held true for DrInC subscores, where across every category except interpersonal, ED men had the highest, ED women had the second highest, and RHC women had the lowest average DrInC scores (Table 2; Figure 2). In the interpersonal subscore, the difference in average DrInC scores was not statistically significant. However, in all subcategories except social responsibility and impulse control, ED women had the highest range of DrInC scores (Figure 2).

**Figure 2:**
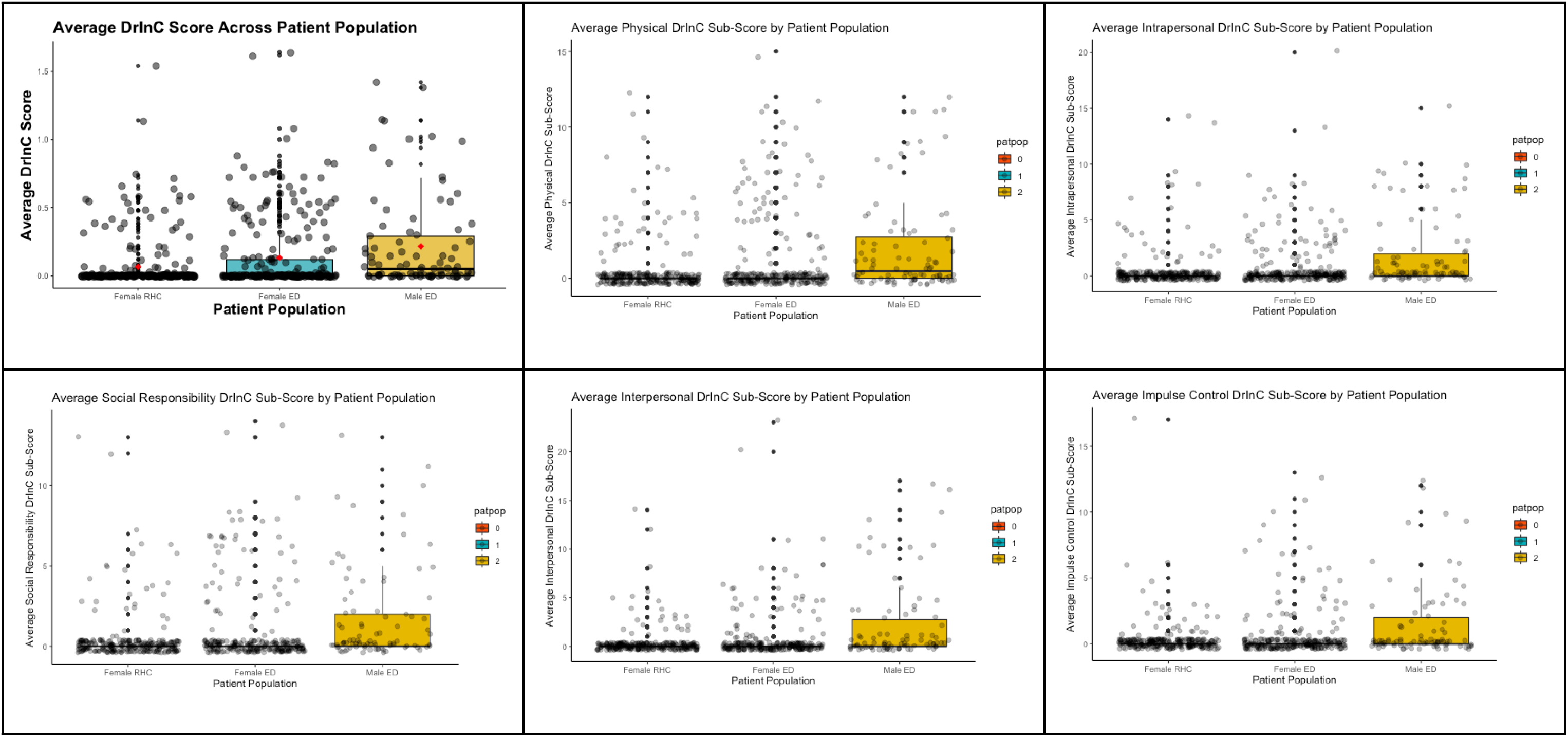
Box-Plot Distribution of DrInC Scores and Subscores across Patient Populations

**Table 2:** Reported Average DrInC Scores and Subscores across Patient Populations.

|  | **RHC Women**<br>**(n =276)** | **ED Women**<br>**(n = 275)** | **ED Men**<br>**(n = 122)** | **Total Population**<br>**(n = 678)** |
| --- | --- | --- | --- | --- |
| **Overall DrInC Score, *missing: 17*** | 5.47 (9.33, 75) | 9.11 (13.1, 80) | 16.4 (19.6, 88) | 9.00 (13.9, ) |
| **Physical Subscore, *missing: 17*** | 0.598 (1.73, 12) | 1.39 (2.75, 15) | 2.54 (3.37, 12) | 1.28 (2.62, 15) |
| **Intrapersonal Subscore, *missing: 17*** | 0.542 (1.87, 14) | 0.996 (2.34, 20) | 2.86 (3.97, 17) | 1.15 (2.68, 20) |
| **Social Responsibility Subscore, *missing: 17*** | 0.517 (1.74, 13) | 1.15 (2.53, 14) | 2.53 (3.77, 17) | 1.15 (2.66, 17) |
| **Interpersonal Subscore, *missing: 17*** | 0.391 (1.51, 14) | 0.993 (2.67, 23) | 3.02 (4.42, 17) | 1.12 (2.91, 23) |
| **Impulse Control Subscore, *missing: 17*** | 0.336 (1.35, 17) | 0.695 (1.89, 13) | 1.99 (3.34, 17) | 0.796 (2.17, 17) |
| Data are presented as mean (SD, range) for quantitative variables, and count (%) for qualitative variables. |  |  |  |  |

### Qualitative

Nineteen individuals (RHC women, n = 5; ED women, n = 5; ED men, n = 9) participated in IDIs (Table 3). The ages of IDI participants spanned from 20 to 70 years, with a mix of education, income, and alcohol intake levels represented.

**Table 3:**
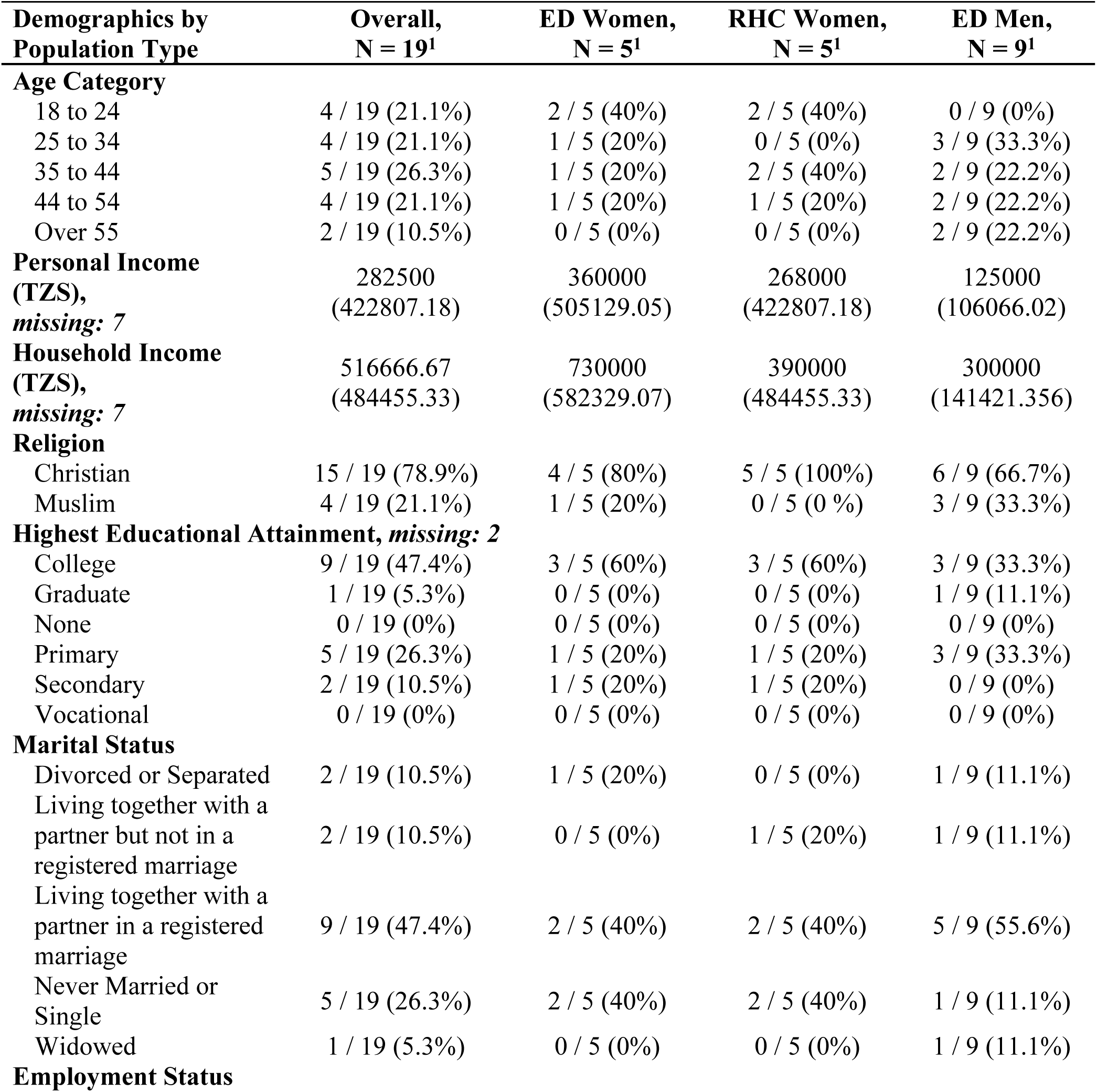
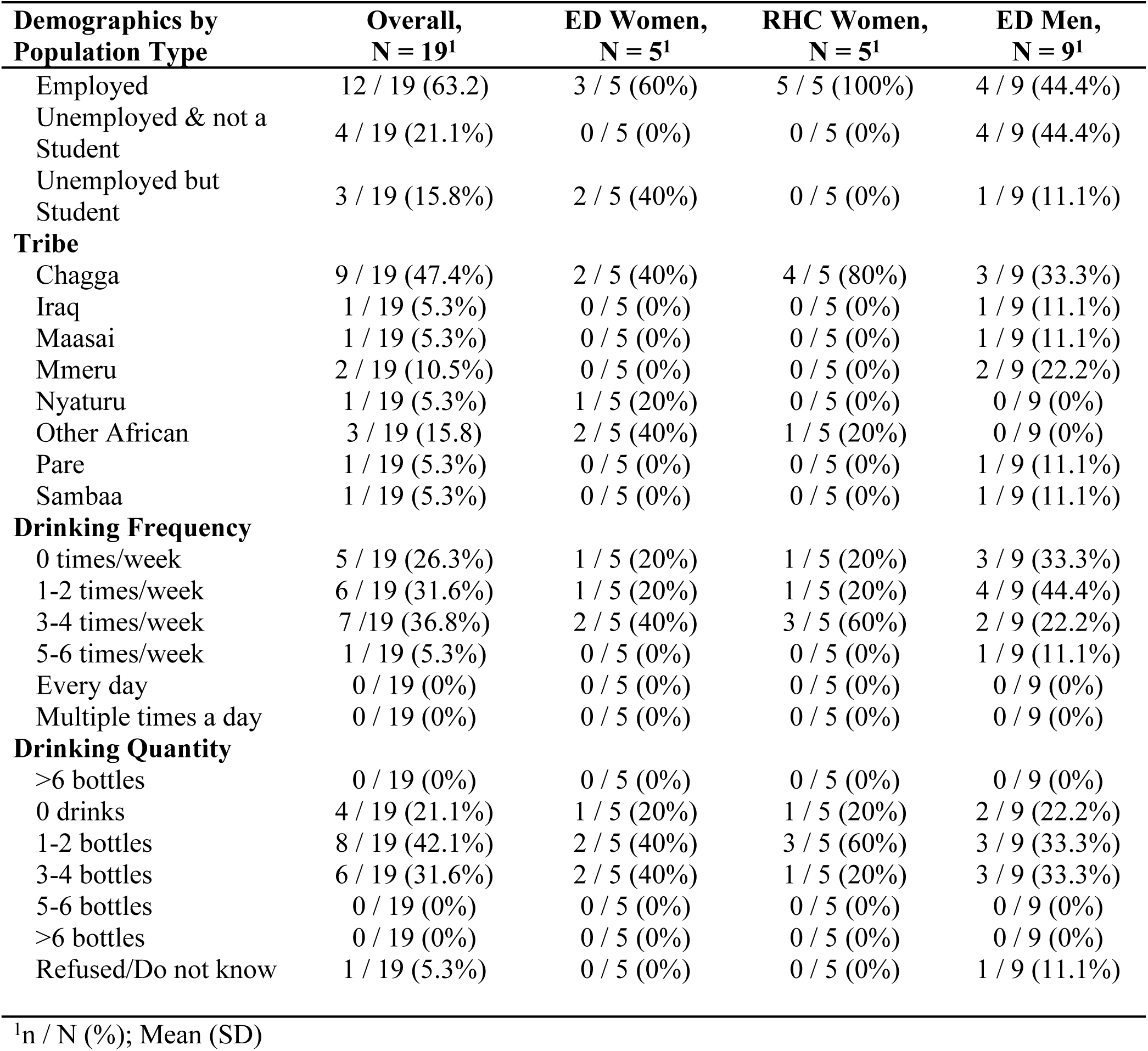
Interviewee Demographics.

Approximately a third of participants held positive views or remarked that they generally benefited from alcohol, almost half said these views and experiences were overall negative, and the remaining quarter of participants stated that it depended. Two main themes and several sub-themes related to community perceptions and the impact of alcohol emerged from the IDIs. The two main themes were Alcohol’s perceived harms and Alcohol’s perceived benefits (Table 4). Participants noted the ways they felt alcohol harmed either them or their community, which included a failure to fulfill their responsibilities, stigma, sexual assault, increased conflict, chronic health issues, and financial instability. On the other hand, several examples of how alcohol was seen as beneficial within the community were mentioned, including fostering economic growth, creating an outlet to uphold cultural traditions, and instilling more social unity.

**Table 4:** Qualitative Themes and Sub-Themes.

| Themes | Sub-Themes |
| --- | --- |
| Alcohol's Perceived Harms | Inability to Fulfill Familial Responsibilities<br>Stigma and Discrimination<br>Sexual Consequences<br>Interpersonal Conflict and Violence<br>Injuries and Chronic Disease<br>Financial Instability |
| Alcohol's Perceived Benefits | Economic Growth<br>Upholding Cultural Practices<br>Social Unity |

To preface this data, the communities’ perceptions and expectations of alcohol use explored in this results section is the amalgamation of general themes and trends from 19 participants. While these findings have been grouped for clarity, it is important to note that there was a significant amount of variability in responses and opinions, especially across gender, “*various religions, clans, and tribes” (IDI #3, Female)*. As one participant explains:

> “The community is the combination of many people with different ideologies about life; there is no way that my perception will be the same with everybody in the community. It’s because we have been born and raised in different families and environments and everyone came up with his/her own life basics, so according to how one came across alcohol it is how the perception will be built..” (IDI #19, Female)

### Alcohol’s Perceived Harms

As part of their interview, IDI participants were asked how alcohol use negatively impacted their community. Respondents stated that alcohol leads to stigma, sexual violence, risky sexual behaviors, and an inability of community members to uphold individual responsibilities. Most also saw it as an instigator of physical, verbal, and emotional conflict among family members, and believed it contributed to long-term health issues and financial instability.

> “What I can say is the society do not have positive expectation about alcohol use, because we see families disintegrated, people fall sick as a result of alcohol and many people at first drink but are able to work and carry on their daily activities and routine but later on they fail to carry on tasks because alcohol makes them weak.” (IDI #3, Female)

#### Inability to Fulfill Family Responsibilities

Most participants mentioned that alcohol was an inhibitor in one’s ability to fulfill their responsibilities to their family. Those who drank in excess would “*forget [their] responsibilities*” (IDI #11, Male), become too “*weak*” (IDI #3, Female), “*lazy*” (IDI #19, Female), or simply, “*fail to perform their daily duties*” (IDI #6, Female). One participant elaborates on their own experience:

> “I see alcohol is not such a good thing…It can make one forgetting everything about his/her family, even children can go to bed hungry without you having an idea because you’re busy drinking. Children left home by drunkard parents can even experience assaults of different kinds just because the parents are not there to protect them…I advice my fellow parents and those who have families to care for to stop taking alcohol” (IDI #7, Female)

#### Stigma and Discrimination

All interviewees noted alcohol-related stigma to be present to some extent in their communities, but to varying degrees and concentrated in certain circumstances. One participant reported that they “*haven’t seen any stigma among men or women*…*unless if after drinking you cause problems like fighting or abusing other people*” (IDI#12, Male). As another explains, stigma could also arise when one fails to accomplish preset responsibilities or drinks unsafe alcoholic beverages:

> “Alcohol intake in Tanzania is like a ritual. Anyone can drink. The stigma arises when someone fails to control their drinking habit, like when someone drinks all the time and fails to fulfill his or her obligations, or if someone drinks those homemade beers (local brews) that are not tested and not standardized by the bureau of standards. People who drink Gongo and Dadii are really stigmatized in the community. Because once you drink those, you may even fail to control your bowels, which leads to shame and embarrassment” (IDI #2, Male).

In addition to the quality of liquor, another notable facet of alcohol-related stigma is its interaction with gender. Participants noted that “*women who drink are stigmatized more than men*” (IDI #7, Female) and that their “*community strongly hate women who drinks alcohol”* (IDI #19, Male). This arose in part because of aforementioned gender roles, where because “*women stay with the children most of the time”* (IDI #19, Male) there is concern that their alcohol habits will have greater negative repercussions on their children.

#### Sexual Consequences

Interviewees reported that alcohol use caused or contributed to significant sexual consequences uniquely experienced by each gender. The first of these was the increased risk of acquiring a sexually transmitted disease – *“when you drink too much it can put you a risky of even practice unsafe sexual intercourse”* (IDI #4, Male), which generally stemmed from having more sexual partners:

> “Men who drink lacks discipline, they don’t respect themselves or their wives, they engage in sexual immorality without considering the fact that they are aged, married and with families.” (IDI #7, Female)

A couple of men and women participants noted that alcohol could hinder their ability to perform sexual acts –

> “A person cannot perform sexual activity well as before, to men they may encounter erectile dysfunction and women may also not be able to involve and perform sexual activity properly. Alcohol can make a person really desire the sexual intimacy but just not able to perform it because of the effect of alcohol in the brain.” (IDI #16, Female)

This, though, was only mentioned by a few participants and stands in sharp contrast to the common theme of alcohol-related sexual assault. This final sexual consequence was described almost exclusively as men being violent against women – “*there is someone I know she was invited to a man house to drink alcohol it was her first time to drink alcohol she got drunk the man raped her*” (IDI #9, Female). Participants disagreed on whether or not men incurred sexual assault when drinking, one saying *“men…cannot experience sexual violence because it is not a common practice in Tanzania”* (IDI #2, Male), while another reported that *“men…when they are drunk, they have been sexually abused by the fellow men”* (IDI #13, Female).

#### Interpersonal Conflict and Violence

Sexual assault was not the only product of the increased violent and aggressive behavior that alcohol use incited. Interpersonal conflict and violence was a theme noted by three-quarters of participants. IDI #19 states,

> “Alcohol causes misunderstandings and fights in society, I have witnessed many times that people get drunk and start to say abusive words, become very aggressive and arrogant.” (IDI #19, Female)

Respondents described how alcohol use instigates emotional and physical conflict and distress within the family – “*I know many families that when the father is drunk, the mother and children become so anxious and afraid to sit with him closely. The most common violence is physical and emotional abuse*” (IDI #17, Male). As seen in this and the next quote, aggression was described as being more common in men – *“it’s very difficult to see women fighting after drinking. But men are always aggressive and violent”* (IDI #3, Female), with the ensuing conflict leading to family disintegration.

Moreover, several participants (4 of the 19 participants) discuss how a child’s mental health is significantly affected by witnessing their parents “*fighting and shoutin*g” (INT#12, Male), a form of emotional conflict, due to alcohol consumption. As a result, many pre-existing interpersonal relationships were harmed or broken – *“I have witnessed many times that people get drunk and start to say abusive words to one another …I can say that alcohol can truly bring people together but can also destroy people’s relationship”* (IDI #19, Female).

#### Injuries and Chronic Disease

While all the categories of harms thus far discussed can lead to significant physical, mental, and emotional problems, a plethora of other long-term health issues stemming from alcohol use like injuries and road traffic incidents, illness and chronic disease were also discussed by most participants.

> “Alcohol can lead to organ damage like liver so there is a lot of complications…I had a friend of mine who was drinking every day, and he was not eating well so he ends up dying. Because of alcohol intoxication. But before that he was having anemia (low blood), low body weight, and muscle wasting” (IDI #12, Male)

Health issues that were attributed to alcohol use by participants included the following: liver cirrhosis, cancer, kidney failure, “*puffy cheek*” (water retention resulting from alcohol use), neurological insults, and even death. Some participants even recount how local brews —which are “*not tested and not safe*” *(IDI #4, Male)* and have been characterized as “*strong alcohol*” *(IDI #8, Male)*— may incite even greater health consequences in regular consumers.

#### Financial Instability

Lastly, almost half of the participants mentioned economic loss and financial instability, encouraged via a variety of different mechanisms, as a major source of personal and community harm. First, participants proposed that economic loss and financial instability seem to arise from the perceived difficulty of drinkers to find or maintain employment. That is, participants agreed that employed individuals who drink experience a loss of productivity. IDI #17, for example, stated, *“Alcohol-related behaviors may lower the individual performance at (the) workplace which leads to inadequate or inefficient production” (IDI #17, Male)*.

In addition, if an individual spends *“a lot of money buying alcohol”* (IDI #12, Male) for themselves, this can take a toll on personal savings –*“I lost my business because of drinking too much”* (IDI #12, Male). One participant describes the monetary loss alcohol-related spending could cause:

> “Alcohol affected my uncle so badly, he was so rich, and helped people in his family and relatives but once he became addicted with alcohol everything turns to zero. He lost everything, even closer friends. He is unable to take care of his family even paying school fees for his children!” (IDI# 5, Male)

As men especially were seen to be the primary breadwinners for their families, a father’s excess spending on alcohol could “*decrease in family income”* (IDI #17, Male). This created negative repercussions for a man’s wife and children and permanently cripple their financial holdings:

> “Alcohol is a reason for many families here to fall into extreme poverty because of men as family providers spend a lot of resources in alcohol while they just earn little to help their families survive as most of them work as drivers of tricycles and motorcycles. So, when a man has no control of his drinking habits and spends all of his income into drinking and definitely the family get into a vicious cycle of poverty.” (INT #19)

While increased alcohol consumption was seen as contributing to personal or familial financial distress for some, as will be discussed in the next section, several participants noted a financial benefit to increased alcohol use for sellers or the government.

### Alcohol’s Perceived Benefits

When asked how alcohol has positively impacted their community, a third (only one of which was female) commented on the economic gain in alcohol production and sales, and half of the participants commonly mentioned social and/or cultural benefits.

#### Economic Growth

In contrast to the last sub-theme presented, IDI #2 described the monetary benefit alcohol brought to his community, saying it

> “contributes to the economic growth of an individual who is selling, a family or a national income. The government collects taxes from the brewing industry, which helps to increase national income” (IDI# 2, Male).

Interestingly though, while the government benefited financially from alcohol sales, several respondents discussed how *“local brew”* (IDI #4, Male) is often not effectively tested for alcohol content levels and results in harmful consequences for their community. One participant explained this lack of government monitoring:

> “The community views [alcohol] as a burden promoted by the government, it is the government that legalized it and allow people to manufacture alcohol. And sometimes the government fails to monitor these industries and see what kind of alcohol they are producing. It is very unfortunate that some of the businessmen are not faithful and manufacture alcohol which are very strong and damage people’s brain.” (IDI #12, Male)

Importantly, while the production of alcohol was seen to bring economic benefit for some, even this perceived boon carried with it the potential for harm to community members.

#### Upholding Cultural Practices

As mentioned in the first quote in this manuscript, alcohol use was tied to cultural and tribal events – *“Many people consider alcohol to be part of their social culture and way of life”* (IDI #2, Male). Some respondents found it appropriate to consume alcohol at cultural events or special occasions, using alcohol during rituals as a way to “*signify respect and honor to our ancestors*” *(IDI #2, Male)* and “*honor their ancestors’ way of living, social customs that have been practiced for generations and generations*” *(IDI #3, Female)*. One woman summarized these sentiments:

> “There are social events that cannot be done without alcohol, for example, if you go to pay for dowry you have to bring alcohol to the event and even in some rituals, they must brew alcohol during the ritual service. So, there’s a positive way alcohol affects our society as a whole…many people drink alcohol to spend time together and enjoy” (IDI #9, Female)

#### Social Unity

Finally, alcohol’s positive influence on the social aspect of community life was a common theme in IDIs – *“alcohol helps people interact and have time to discuss different issues…it strengthens unity among people” (IDI #4, Male)* and

> “[Alcohol] helps to bring people together in unity. You may find friends or just random people sitting together, making stories about a thing or two and celebrating their achievement over a glass of beer. In my society, you cannot just gather around grown-up people to discuss about something and just give them soda or juice, you must offer them alcohol so that they will listen to you properly”(IDI #19, Female).

> “The good results of alcohol, to me personally it has made my social life very great and I have been meeting a lot of people when I go for a drink, for my friends as we enjoy drinks together and talk about our lives but I have also met important and influential people to discuss important matters in business and work as we enjoy 1 or 2 glass of beer” (IDI #10, Female)

Not only was alcohol use noted as bringing people together, but alcohol was also mentioned as having a key role in resolving social conflict and social gatherings:

> “If you have misunderstandings with friends or family members, and you want to resolve those disputes…you need to offer them alcohol while you talk! You can’t offer them soft drinks! Even when you want to get married, you need to buy a lot of alcohol for elderly people to drink and enjoy” (IDI #2, Male).

## Discussion

This paper explored the perceptions of alcohol-related consequences through the lens of KCMC ED and RHC patients. This analysis is the first of its kind to provide a holistic overview of the harms and benefits of alcohol as perceived by patients in Moshi, Tanzania. Existing literature on Moshi has described the scope of alcohol use in this region (26,28,32,47), examined the determinants of use (48), and explored specific consequences related to alcohol, such as drunk driving (49) and stigma (29,33). Perceptions of alcohol use have been explored (23), but without looking at alcohol-related consequences and exclusively from the perspectives of injury patients. Our paper takes the existing data one step further by providing a comprehensive view of perceived alcohol-related impacts on individuals, their families, and their communities to allow for a clearer picture of alcohol’s role in this context. Our results show that individuals and their families experienced alcohol-related social harms like stigma and interpersonal conflict; however, alcohol was also thought to foster unity at the broader community level. From a social dimension, while patients perceived alcohol to have some level of financial benefit on the community and government level through sales and taxes, substantial economic harm from excess spending on alcohol arose for the individuals and families of heavy users. Finally, no health benefits were linked to alcohol consumption but a slew of physical harms were mentioned by participants.

In this analysis, we found that participants saw both positive and negative impacts of alcohol use at the individual, interpersonal, and community levels, including the perceived benefit of social unity and upholding traditional customs contrasted by the harm of greater conflict and stigma. One notable stipulation to these results, though, is that while ED men had the highest average scores on the DrInC social responsibility category, stigma was found to more severely affect women. Looking first at the social consequences to the individual, previous Moshi-based studies have identified alcohol-related stigma (33,50) and a disproportionate stigma against women (29), however greater exploration on this topic is needed as these data sets were limited to ED injury patients. At the interpersonal level, alcohol use was purported to foster social connections but at the same time had the potential to generate conflict, going as far as to destroy romantic and family relationships. Though the first of these findings is not well-supported by academic literature, alcohol has been previously noted to be a mediator in conflict and intimate partner violence both globally and regionally (51–54). While both social benefits and harms were noted by participants, only the latter is currently supported in scholarly publications. However, understanding the perceived positive associations of alcohol use can help better contextualize its ubiquitous usage in Moshi and pinpoint focus areas for future alcohol-reduction-focused interventions.

Similar to the social implications of alcohol use, we found that alcohol consumption can have important financial consequences at the individual and country levels. IDI participants described the decreased household income that arose from excessive spending on alcohol. This is supported by other previously published study data where we found nearly 40% of male ED patients spent between 10 - 46% of overall average monthly household income on alcohol alone (39,55). While women are spending the least amount on alcohol overall, previous data showed that women consume less alcohol but also experience limited autonomy when it comes to the use of household funds. Other important economic-based findings from this analysis are the impacts at broader community and government levels. IDI participants believed that high rates of consumption were advantageous for sellers of alcohol and rising national excise taxes on alcohol products contributed to increasing revenue for the government of Tanzania. However, specifics of how this additional revenue was used to benefit Tanzanians were not mentioned. Currently, while there is a lack of literature outlining the broader financial implications of alcohol use within the East African context. Matzopoulos et al. estimate that within South Africa, the tangible and intangible costs incurred from harmful alcohol consumption comprise between 10-12% of the country’s total GDP and outweigh the economic benefits associated with alcohol taxes and sales (56). Further, research has suggested the inverse relationship between alcohol consumption and labor force participation globally (57). This contributes to larger macroeconomic implications, including losses in economic productivity and development. Overall, our data suggest that financial benefits from alcohol use were perceived to only exist for sellers of alcohol and at the government level. For individuals and the broader community, participants almost exclusively noted the negative monetary impacts of alcohol use.

Along with the social and financial consequences of alcohol consumption, we noted several physical harms stemming from alcohol use. ED men were seen to experience a more diverse range of physical consequences than ED and RHC women as illustrated by our DrInC physical harm scores. We found in IDI data that perceived alcohol-related physical harms include increases in sexual assault, violence, risky sexual behaviors, and chronic health issues, with no mention of any physical benefit to themselves or their communities. This last finding echoes a recent statement made by the World Health Organization (WHO) stating that any level of drinking is harmful to physical health (58,59). Elaborating on these harms, the physical consequences of alcohol use mentioned by our IDI participants have been noted in other Africa-based studies. In sub-Saharan Africa, harmful alcohol use has been previously associated with increased road traffic injuries, drunk driving, and other injurious behaviors (32,49,60,61). In addition, studies in Uganda and South Africa have found an association between sexual and intimate partner violence and alcohol consumption (62,63). Last, our IDI data also echoes the WHO and the Center for Disease Control’s growing concern over the chronic health consequences of excess consumption such as liver and kidney failure (64). Effective alcohol-reduction programs stand to reduce not only these negative health effects associated with alcohol but also violent behaviors that incite injuries. Walton et al. and Ward et al. found, respectively, that interventions aiming to lower alcohol intake subsequently reduce aggression (65,66). Given the notable physical risk that alcohol poses, there is a need to address alcohol use behaviors in the Moshi area. Additionally, as research has shown that interventions are more effective when culturally adapted to specific populations (67,68), alcohol-reduction-related programs and policies implemented in Moshi should encompass the specific socio-cultural nuances of the region.

In considering the social, financial, and physical impacts of alcohol use, one final note is that while broader community benefits like social unity, the upholding of traditions, and increased government revenue were raised, participants identified few personal benefits of alcohol use. This raises the question of if alcohol truly has benefits in the community outside of historic incorporation into cultural and social events. More research exploring alcohol’s impact at the personal versus community levels are thus recommended.

### Limitations and Directions for Future Research

Limitations for this analysis are present primarily within the quantitative data set and the DrInC tool. First, the rate of patients who screen failed or declined survey participation was not originally collected. This has an impact on the generalizability of our quantitative data to the ED and RHC patient populations at large. Additionally, this data was collected in the midst of the COVID-19 pandemic. As such, for the safety of our Tanzanian research staff, those who presented to the ED for COVID-19-related symptoms were not approached for study participation. While necessary to take these safety precautions, this also limited our ability to reach the full expected patient population. In both these cases, however, because of the large sample size and well-enforced sampling strategy, we still hold our data to be representative of the patient population. For DrInC, while this tool helps measure the range of consequences one might experience from alcohol use, it does not provide information on how severe these consequences are to the individual, which limits our understanding of alcohol’s true impact. Finally, several important gender-specific consequences of alcohol use arose in this analysis, highlighting the need for further exploration of how alcohol differentially impacts men and women in this region. Even with these limitations, our paper offers one of the first published assessments of community perceptions of alcohol use in sub-Saharan Africa, offering insight into how alcohol is viewed both positively and negatively by patients and community members. Our work highlights the need to consider social, financial, and cultural elements when addressing alcohol use disorders.

## Conclusion

Alcohol was perceived to have both positive and negative impacts on patients and their communities. Perceived harms included stigma, sexual harm, interpersonal conflict, injuries and disease, and financial instability. These harms stand in contrast to the mentioned benefits of increased social unity, upholding traditional practices, and economic growth. Further, men were found to experience a wider range of alcohol-related consequences than women. Our results allow for a sharper picture of alcohol’s role in this community and point to the need to incorporate the sociocultural, financial, and physical implications of alcohol use in programming and treatment related to alcohol use disorders. These findings can help identify focus areas for future programs aiming to reduce alcohol misuse in this area.

## Author contributions

Conceptualization: AMP, CAS, BTM, BM

Methodology: AMP, CAS, JRNV

Formal analysis and investigation: CAS, BTM, BM

Data Collection: MMi, JK, YS, AMP

Writing - original draft preparation: AMP, MB, KW, MM, WN

Writing - review and editing: SR, AMP, MB, KW, MM, WN, CAS

Funding acquisition: AMP, CAS

Supervision: CAS, SR, JRNV, BTM

## Data Availability

Data are only available upon reasonable request, as participants did not consent to public data publishing, and data transfer requires a written agreement approved by Kilimanjaro Christian Medical Centre Ethics Committee and the National Institute for Medical Research (Tanzania). Data inquiries can be sent to Gwamaka W. Nselela at.

